# Public perspectives on social distancing and other protective measures in Europe: a cross-sectional survey study during the COVID-19 pandemic

**DOI:** 10.1101/2020.04.02.20049676

**Authors:** Karien Meier, Toivo Glatz, Mathijs C Guijt, Marco Piccininni, Merel van der Meulen, Khaled Atmar, Anne-Tess C Jolink, Tobias Kurth, Jessica L Rohmann, Amir H Zamanipoor Najafabadi, on behalf of the COVID-19 Survey Study group

## Abstract

**Objectives:** The extent to which people implement government-issued protective measures is critical in preventing further spread of coronavirus disease 2019 (COVID-19) caused by coronavirus SARS-CoV-2. Our study aimed to evaluate the public belief in the effectiveness of protective measures, the reported implementation of these measures in daily life, and to identify communication channels used to acquire relevant information on COVID-19 in European countries.

**Design:** A cross-sectional online survey available in multiple languages was disseminated on social media starting March 19th, 2020. After five days, we computed descriptive statistics for countries with more than 500 respondents. Each day, we compiled and categorized community containment measures enacted in each country by stringency (stage I-IV). Response collection continued for one week to explore possible dynamics as containment strategies intensified.

**Participants:** In total, 9,796 adults responded, of whom 8,611 resided in the Netherlands (stage III), 604 in Germany (stage III), and 581 in Italy (stage IV). An additional 1,365 respondents completed the survey in the following week.

**Results:** Participants indicated support for governmental measures related to avoiding social gatherings, selective closure of public places, and hand hygiene and respiratory measures (range for all measures: 95.0%-99.7%). Respondents from the Netherlands were less likely to consider a complete social lockdown effective (59.2%), compared to respondents in Germany (76.6%) or Italy (87.2%). Italian residents did not only apply enforced social distancing measures more frequently (range: 90.2%-99.3%, German and Dutch residents: 67.5%-97.0%), but also self-initiated hygienic and social distancing behaviors (range: 36.3%-96.6%, German and Dutch residents: 28.3%-95.7%). Respondents largely reported being sufficiently informed about the COVID-19 outbreak and about behaviors to avoid infection (range across countries: 90.2%-91.1%). Information channels most commonly reported included television (range: 53.0%-82.0%), newspapers (range: 31.0%-63.0%), official health websites (range: 39.0%-54.1%), and social media (range: 40.0%-55.8%). We observed no major changes in answers over time.

**Conclusions:** In European countries, the degree of public belief in the effectiveness of protective measures was high and residents reported to be sufficiently informed by various communication channels. In March 2020, implementation of enacted and self-initiated measures differed between countries and were highest among Italian respondents, who were subjected to the most elaborate measures of social lockdown and greatest COVID-19 burden in Europe.

## INTRODUCTION

The recent pandemic of COVID-19 (coronavirus disease 2019) caused by SARS-CoV-2 (Severe Acute Respiratory Syndrome Coronavirus 2) has infected more than 1,000,000 people worldwide in only a few months’ time and caused more than 51,000 deaths as of April 2nd, 2020[1]. This rapidly spreading virus imposes a tremendous burden on national healthcare systems, as they lack sufficient material and human resources to respond to the rapidly increasing number of patients requiring intensive care[1,2].

Worldwide, public health organizations, as well as national and international government bodies, have suggested systematic implementation of protective, public health measures in an effort to delay the spread of COVID-19[3]. The aim of these measures is to decrease the peak infection rate, while maintaining a high quality of care under finite resources and limited hospital capacities[2,4]. In addition to basic hygienic advice such as regular hand washing, the most important recommendation known to limit and delay the spread of the virus is social (physical) distancing[5,6].

In early March 2020, Europe became the epicenter of the COVID-19 pandemic, with more cases and deaths reported than in all other countries (excluding China) combined[1]. Throughout the course of the month, most European countries progressively implemented community isolation measures to increase social distancing, such as imposing work restrictions and the closure of public places and shops. Italy, the first and most severely affected country in Europe, imposed strict community isolation measures on March 9th and 11th, 2020, enforcing a nationwide quarantine in response to the alarming increase in the number of cases, which posed a serious threat to the capacity of the Italian healthcare system[7].

The extent to which people are informed about and apply the measures advocated by experts and enforced by governments is critical to control the spread of the virus and to optimize patient outcomes during the current COVID-19 pandemic[4,8,9]. The aim of our study was to evaluate public belief in the effectiveness of protective measures, to what extent individuals have implemented these measures in their daily lives, and to identify key communication channels used to acquire information on COVID-19 in European countries. We believe these insights are not only valuable for the ongoing mitigation of the current COVID-19 pandemic, but may also serve to inform governments’ and public health organizations’ information dissemination and infection control strategies for possible future pandemics.

## METHODS

### Design, setting, and participants

The survey instrument used to gather cross-sectional data was compiled by a team of medical students and epidemiologists from the Leiden University Medical Center and the Charité - Universitätsmedizin Berlin. Our initial aim was to collect sufficient data on adults living in Europe, with an emphasis on individuals residing in the Netherlands, Germany, and Italy, however, the survey was also open to residents of other countries. Data collection is still ongoing and is planned to continue for as long as community isolation measures remain in place. In these primary descriptive analyses, we only reported data from countries with at least 500 responses at our first cut-off date, March 23rd, 2020. This date was chosen because several European regional and national governing bodies announced stricter measures around this date. The study was reviewed and granted exempted status from medical ethical approval by the Institutional Review Board of Leiden University Medical Center in The Netherlands (protocol number: N20-037).

### Survey instrument

We selected questions from the validated Flu TElephone Survey Template (FluTEST), which was designed to assess perceptions and behavior during an influenza pandemic [10]. We slightly modified the items to fit the current outbreak context, where necessary and formulated additional questions to assess beliefs in the effectiveness of protective measures [11]. In brief, the survey instrument consisted of 22 items in three sections including: 1) five questions regarding beliefs in the effectiveness of public measures to reduce outspread (e.g. selective closure of places and complete social lockdown), 2) 16 questions on the personal application of protective measures (e.g. social distancing behaviors and hygienic practices), and 3) one question on the three most frequently used sources to acquire information on the COVID-19 outbreak, and one question on the perception whether or not respondents felt sufficiently informed. The full survey is presented in Supplemental Text 1. To allow for stratified interpretation of the results, additional questions captured sociodemographic information about gender, age, household composition, employment status, educational level, country of residence, being a healthcare provider or (bio)medical student, and prevalent chronic medical conditions. Respondents were able to complete the questionnaire only once per device in an effort to reduce potential repeat responses.

The survey was translated into multiple languages by a small panel of native speakers from the original English language version. Due to time constraints, we were unable to formally validate the survey questions in the other languages. The survey went live on March 19th, 2020 in Dutch, English and German. Other languages have been added since initiation (Italian on March 20, 2020; French and Polish on March 21, 2020; Spanish on March 22, 2020; Turkish on March 25, 2020; and Farsi on March 29, 2020; see Supplemental Text 1).

### Procedures

The full survey was initially piloted on a sample of 50 respondents. After minor modifications to the structure and language, the survey was actively disseminated through (social) media channels, such as WhatsApp, Telegram, Facebook, LinkedIn, Instagram, and Twitter, and in professional networks via electronic mailing lists. The survey was further promoted via a number of local and national news websites and radio stations. On the landing page, participants were briefed about the study and only those providing informed consent for participation were guided to the actual 5-minute survey. On the final page, participants were debriefed about the study, and thanked for their contribution.

### Assessment of stages of community containment measures

We extracted community containment measures taken by governments in each country included in this study from national governmental announcements and news articles and compiled daily from March 1st, 2020 onwards. Two independent researchers classified stringency of isolation measures by country in four stages based on the Community Containment Measures guideline originally developed by the Centers for Disease Control and Prevention (CDC) during the SARS outbreak in 2003[12]. Any disagreement was resolved by discussion. The CDC guideline describes seven interventions, which we grouped into four stages to create country-specific timelines for the purposes of this study. Guideline interventions 1 (passive monitoring), 2 and 3 (active monitoring without and with activity restrictions, respectively) were grouped together as Stage I (“Low Impact Containment Measures”), since most countries had already implemented these interventions in early March 2020. Guideline interventions 4 (working quarantine) and 5 (focused measures) were grouped and classified as Stage II (“Focused Measures to Increase Social Distance”), as many countries applied these interventions simultaneously. We designated intervention 6 as Stage III (“Community-Wide Measures to Increase Social Distance”) and intervention 7 as Stage IV (“Widespread Community Quarantine, Including Cordon Sanitaire”). We detailed and justified the daily stage classification by country in a series of timelines (Supplemental Tables 1a-1d; Supplemental Text 2).

**Table 1.** Sociodemographic characteristics of respondents on March 23rd, 2020, by country.

|  | Netherlands<br>(Stage III) | Germany<br>(Stage III) | Italy<br>(Stage IV) | Total |
| --- | --- | --- | --- | --- |
| No. | 8,611 | 604 | 581 | 9,796 |
| **Gender (%)** |  |  |  |  |
| Male | 2,492 (28.9) | 206 (34.1) | 192 (33.1) | 2,890 (29.5) |
| Female | 6,095 (70.8) | 389 (64.4) | 388 (66.8) | 6,872 (70.2) |
| Other | 24 (0.3) | 9 (1.5) | 1 (0.2) | 34 (0.4) |
| **Age, years (%)** |  |  |  |  |
| ≤20 years | 675 (7.8) | 11 (1.8) | 117 (20.1) | 803 (8.2) |
| 21-30 years | 2,867 (33.3) | 222 (36.8) | 189 (32.5) | 3,278 (33.5) |
| 31-40 years | 1,167 (13.6) | 223 (36.9) | 99 (17.0) | 1,489 (15.2) |
| 41-50 years | 1,278 (14.8) | 70 (11.6) | 75 (12.9) | 1,423 (14.5) |
| 51-60 years | 1,558 (18.2) | 47 (7.8) | 73 (12.6) | 1,678 (17.1) |
| 61-70 years | 852 (9.9) | 23 (3.8) | 26 (4.5) | 901 (9.2) |
| > 70 years | 214 (2.5) | 8 (1.3) | 2 (0.3) | 224 (2.3) |
| **Daily activity (%)** |  |  |  |  |
| Paid job | 4,885 (56.7) | 388 (64.2) | 321 (55.3) | 5,594 (57.1) |
| Homemaker/Unemployed | 573 (6.7) | 41 (6.8) | 37 (6.4) | 651 (6.7) |
| Student | 2,272 (26.4) | 131 (21.7) | 199 (34.3) | 2,602 (26.6) |
| Retired | 557 (6.5) | 20 (3.3) | 12 (2.1) | 589 (6.0) |
| Other | 324 (3.8) | 24 (4.0) | 12 (2.1) | 360 (3.7) |
| **Household composition (%)** |  |  |  |  |
| Single | 1,391 (16.2) | 113 (18.7) | 94 (16.2) | 1,598 (16.3) |
| Parent(s) with child(ren) | 2,740 (31.8) | 174 (28.8) | 135 (23.2) | 3,049 (31.1) |
| Living with partner | 2,339 (27.2) | 189 (31.3) | 94 (16.2) | 2,622 (26.8) |
| Shared flat (with roommates) | 1,558 (18.1) | 112 (18.5) | 34 (5.9) | 1,704 (17.4) |
| Other | 583 (6.8) | 16 (2.7) | 224 (38.6) | 823 (8.4) |
| **Highest educational qualification (%)** |  |  |  |  |
| Primary/Secondary | 635 (7.4) | 132 (21.9) | 301 (51.8)* | 1068 (10.9) |
| Tertiary vocational | 1,991 (23.1) | 57 (9.4) |  | 2048 (20.9) |
| Tertiary academic | 5,985 (69.5) | 415 (68.7) | 280 (48.2) | 6,680 (68.2) |
| **Healthcare provider/(bio-)medical student (%)** | 1,572 (18.3) | 123 (20.4) | 48 (8.3) | 1,743 (17.8) |
| **Chronic illness or being in poor medical condition (%)** | 1,528 (17.7) | 103 (17.1) | 64 (11.0) | 1,695 (17.3) |
| **Sources used to acquire information on COVID-19 (%)** |  |  |  |  |
| Television | 6,613 (76.8) | 320 (53.0) | 476 (82.0) | 7,409 (75.6) |
| Newspaper, mobile news application | 5,422 (63.0) | 295 (48.8) | 180 (31.0) | 5,897 (60.2) |
| Social media | 3,441 (40.0) | 277 (45.9) | 324 (55.8) | 4,042 (41.3) |
| Radio | 1,077 (12.5) | 161 (26.7) | 31 (5.3) | 1,269 (13.0) |
| Official health hotlines | 127 (1.5) | 7 (1.2) | 9 (1.6) | 143 (1.5) |
| Official health websites | 3,361 (39.0) | 327 (54.1) | 268 (46.1) | 3,956 (40.4) |
| Healthcare professionals | 381 (4.4) | 26 (4.3) | 44 (7.6) | 451 (4.6) |
| People I speak to on a daily basis | 2,293 (26.6) | 161 (26.7) | 139 (23.9) | 2,593 (26.5) |

*For full questionnaire and wording, see Supplement 1.*
\* Based on differences in the Italian education system, we considered primary, lower secondary school and upper secondary school as “Primary/Secondary” and university degrees as “Tertiary academic”.

### Statistical Analyses

Data collected over a five-day period between March 19th, 2020 and March 23th, 2020 at 11:20 AM (UTC+0), were used in the primary analyses. We present results of the survey items including sociodemographic characteristics using descriptive summary statistics for the countries having more than 500 responses during this primary data collection period (the Netherlands, Germany, and Italy). Nominal variables were described and visualized using frequencies and percentages. We also reported frequencies of missing responses. We present stratified results for the assessed sociodemographic variables only for the Netherlands, as the number of responses was sufficient per individual subgroup. No formal statistical comparisons were made between countries since the primary aim was descriptive and there were no *a priori* testable hypotheses.

As a secondary analysis, we explored changes in responses for items about the beliefs in the effectiveness of these measures and their implementation over time. As for items about implementation of protective measures, we reported the proportion of positive answers (“Yes”) out of all responses, excluding responses indicating the question was not applicable to their situation. Similarly, for items about the belief in effectiveness of these measures, we considered the proportion of positive answers (“Probably true”). To easily visualize the change over time, we modeled the proportions for each item and for each country separately, using generalized additive models with time as the independent variable, using a shrinkage version of cubic splines with three knots. In addition, we computed and presented visualizations of the differences in proportion between the responses recorded during the primary data collection period and the weeklong extension only for the Netherlands.

Data management, analyses and visualizations were conducted using Stata 16.1 (StatCorp LP, College Station, TX) software and R 3.6.3 / RStudio 1.2.

## RESULTS

Between March 19th and 23rd, 2020, a total of 9,796 respondents completed the survey. Three countries met our study inclusion criteria of more than 500 respondents; the Netherlands (n=8,611), Germany (n=604), and Italy (n=581) (see flowchart: Figure 1). During this primary data collection period, the containment measures in the Netherlands and Germany met criteria for Stage III classification (“Community-Wide Measures to Increase Social Distance”), and those in Italy met Stage IV criteria (“Widespread Community Quarantine”).

**Figure 1.**
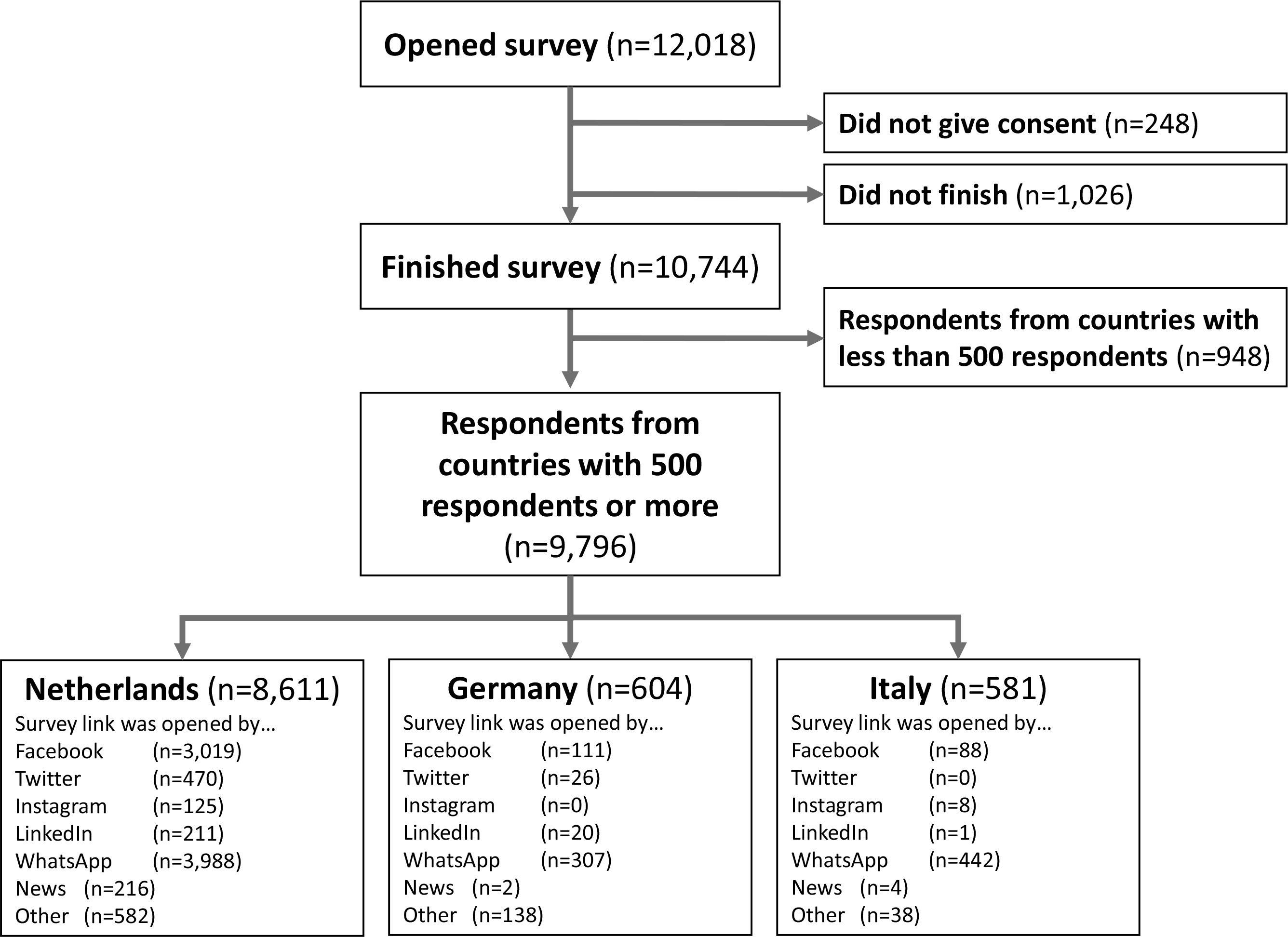
Flow chart of respondents in survey, on March 23rd, 2020.

Most respondents opened the survey link via WhatsApp or Facebook (Figure 1). Approximately two-thirds of respondents were female and one-third of respondents were aged 21-30 years old (Table 1). The majority of respondents had a paid job (57.1%) and many had tertiary academic degrees (68.2%) Approximately 18% of respondents were healthcare providers or (bio)medical students. Less than one-fifth of respondents reported suffering from a chronic illness or being in poor medical condition (17.3%). Descriptive sociodemographic characteristics stratified by country of residence are presented in Table 1.

### Sources used to acquire information on the COVID-19 outbreak

Among respondents living in the Netherlands, Germany, or Italy, the most frequently used sources to obtain relevant information included television (e.g. news, range: 53.0%-82.0%), newspapers or news applications (range: 31.0%-63.0%), social media (e.g. Facebook and Twitter, range: 40.0%-55.8%), and official health websites (range: 39.0%-54.1%). Other people (e.g. family, friends and colleagues, range: 23.9%-26.7%) and radio were reported less frequently as sources of information (range: 5.3%-26.7%). In all three countries, healthcare professionals (range: 4.3%-7.6%) and official health hotlines (range: 1.2%- 1.6%) were the least frequently reported sources of information (Table 1). Almost all respondents living in these three countries reported being sufficiently informed about the current COVID-19 outbreak and what they could do to prevent an infection (range: 90.2%-91.1%; Table 2).

**Table 2.** Being informed about and belief in the effectiveness of policy recommendations during the COVID-19 pandemic on March 23rd, 2020, by country.

|  |  | Netherlands<br>(Stage III) | Germany<br>(Stage III) | Italy<br>(Stage IV) |
| --- | --- | --- | --- | --- |
| No. |  | 8,611 | 604 | 581 |
| **Have been sufficiently informed (%)** | Probably true | 7,839 (91.0) | 545 (90.2) | 529 (91.1) |
|  | Probably false | 271 (3.2) | 23 (3.8) | 29 (5.0) |
|  | Not sure | 476 (5.5) | 33 (5.5) | 21 (3.6) |
|  | No opinion | 25 (0.3) | 3 (0.5) | 2 (0.3) |
| **Belief in effectiveness of recommendations (%)** |  |  |  |  |
| Avoid social gatherings | Probably true | 8,479 (98.9) | 594 (98.5) | 576 (99.7) |
|  | Probably false | 60 (0.7) | 4 (0.7) | 2 (0.4) |
|  | Don't know | 31 (0.4) | 5 (0.8) | 0 (0.0) |
|  | Missing | 41 | 1 | 3 |
| Selective closure of public places/locations | Probably true | 8,158 (95.3) | 586 (97.2) | 568 (98.1) |
|  | Probably false | 234 (2.7) | 13 (2.2) | 11 (1.9) |
|  | Don't know | 172 (2.0) | 4 (0.7) | 0 (0.0) |
|  | Missing | 47 | 1 | 2 |
| Implementation of hand hygiene measures | Probably true | 8,240 (96.0) | 589 (98.2) | 567 (98.1) |
|  | Probably false | 169 (2.0) | 4 (0.7) | 8 (1.4) |
|  | Don't know | 169 (2.0) | 7 (1.2) | 3 (0.5) |
|  | Missing | 33 | 4 | 3 |
| Implementation of respiratory measures | Probably true | 8,139 (95.0) | 583 (97.3) | 569 (99.0) |
|  | Probably false | 230 (2.7) | 9 (1.5) | 4 (0.7) |
|  | Don't know | 196 (2.3) | 7 (1.2) | 2 (0.4) |
|  | Missing | 46 | 5 | 6 |
| Complete social lockdown/isolation | Probably true | 5,063 (59.2) | 458 (76.6) | 495 (87.2) |
|  | Probably false | 1,983 (23.2) | 72 (12.0) | 29 (5.1) |
|  | Don't know | 1,500 (17.6) | 68 (11.4) | 44 (7.8) |
|  | Missing | 65 | 6 | 13 |
| *For full questionnaire and wording, see Supplement I.* |  |  |  |  |
| *Response percentages may not add up to 100% due to rounding.* |  |  |  |  |

### Belief in the effectiveness of measures to reduce outspread

The majority of respondents believed that avoiding social gatherings, selective closure of public places and locations, hand hygiene measures, and respiratory measures were effective ways to prevent further spread of COVID-19 (range for all measures in all three countries: 95.0%-99.7%; Table 2). Only 59.2% of respondents in the Netherlands perceived a complete social lockdown or isolation measures as effective, compared with 76.6% of respondents from Germany and 87.2% from Italy (Figure 2 and Table 2).

**Figure 2.**
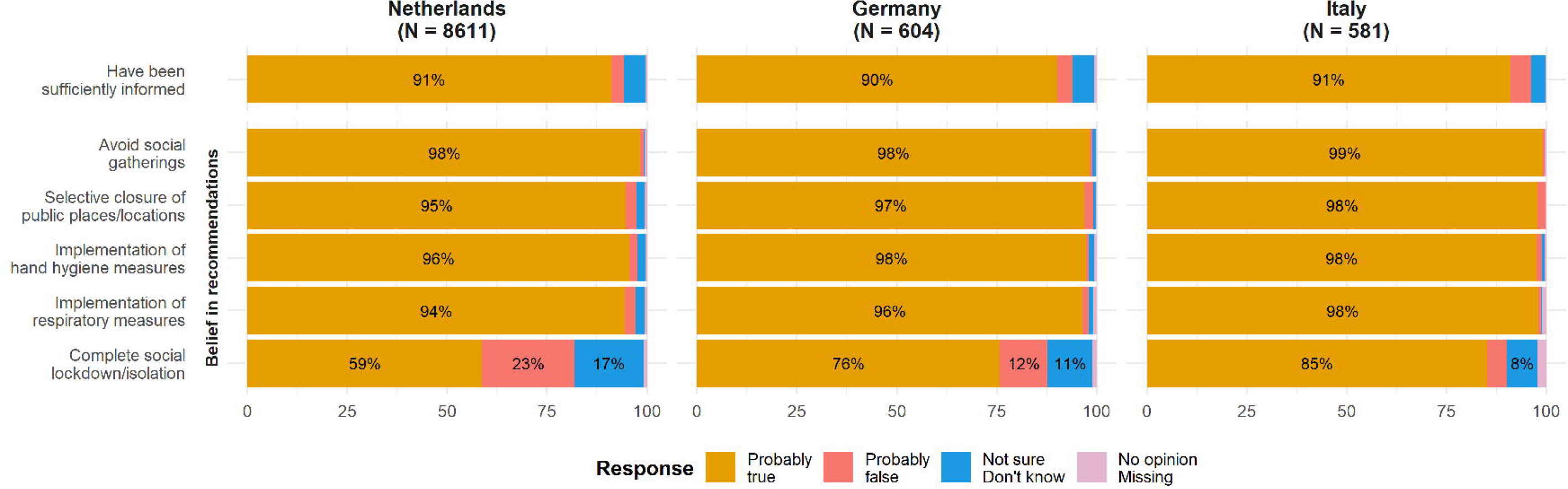
Being informed about and belief in the effectiveness of policy recommendations during the COVID-19 pandemic on March 23rd, 2020, by country. Note: Response percentages are rounded and may not add up to 100%. Percentages below 5% omitted.

### Individual implementation of protective measures

For all items, the percentages reported in the text and Table 3 excluded respondents to whom the item did not apply, which was especially important in the interpretation of three items (keeping children at home before any mandates were put in place, range: 41.0%-75.1%; reducing the use of public transport, range: 1.9%-28.5%; and going to school/university/work, range: 2.4%-14.9%). With regard to personal protective behaviors, a high number of respondents from the Netherlands and Germany reported to have washed their hands with soap and water more often than usual (range: 95.0%-95.7%). In general, respondents from Italy reported applying all proposed personal protective behaviors more often than those from the Netherlands or Germany, except for following a healthy diet or using vitamin supplements (36.3%, Netherlands 54.5%, Germany 54.4%) (Figure 3 and Table 3).

**Table 3.** Individual implementation of protective measures in response to COVID-19 pandemic on March 23rd, 2020, by country.

|  | Netherlands<br>(Stage III) |  |  |  | Germany<br>(Stage III) |  |  |  | Italy<br>(Stage IV) |  |  |  |
| --- | --- | --- | --- | --- | --- | --- | --- | --- | --- | --- | --- | --- |
| No. | 8,611 |  |  |  | 604 |  |  |  | 581 |  |  |  |
| Personal protective behaviors (%) | Y | N | NS | NA | Y | N | NS | NA | Y | N | NS | NA |
| Cleaned or disinfected things you might touch more often than usual | 4,083<br>(47.8) | 3,753<br>(43.9) | 713<br>(8.3) | 62 | 292<br>(48.9) | 264<br>(44.2) | 41<br>(6.9) | 7 | 423<br>(73.2) | 123<br>(21.3) | 32<br>(5.5) | 3 |
| Carried sanitizing hand gel with you when out and about | 2,385<br>(28.3) | 5,871<br>(69.6) | 181<br>(2.2) | 174 | 277<br>(47.0) | 302<br>(51.2) | 11<br>(1.9) | 14 | 424<br>(73.5) | 147<br>(25.5) | 6<br>(1.0) | 4 |
| Used sanitizing hand gel to clean your hands, more often than usual | 4,681<br>(55.3) | 3,601<br>(42.5) | 186<br>(2.2) | 143 | 424<br>(71.5) | 157<br>(26.5) | 12<br>(2.0) | 11 | 442<br>(76.6) | 125<br>(21.7) | 10<br>(1.7) | 4 |
| Reduced the amount you touch your eyes, nose and/or mouth | 4,208<br>(49.0) | 2,491<br>(29.0) | 1,889<br>(22.0) | 23 | 374<br>(62.2) | 87<br>(14.5) | 140<br>(23.3) | 3 | 439<br>(75.7) | 77<br>(13.3) | 64<br>(11.0) | 1 |
| Followed a healthy diet or took vitamin supplements | 4,548<br>(54.5) | 3,429<br>(41.1) | 370<br>(4.4) | 264 | 317<br>(54.4) | 226<br>(38.8) | 40<br>(6.9) | 21 | 205<br>(36.3) | 308<br>(54.5) | 52<br>(9.2) | 16 |
| Usually carried tissues with you when out and about | 4,399<br>(52.4) | 3,833<br>(45.7) | 163<br>(1.9) | 216 | 400<br>(69.7) | 157<br>(27.4) | 17<br>(3.0) | 30 | 472<br>(83.0) | 83<br>(14.6) | 14<br>(2.5) | 12 |
| Usually used tissues when sneezing or coughing | 4,542<br>(62.0) | 2,551<br>(34.8) | 232<br>(3.2) | 1,286 | 335<br>(62.5) | 169<br>(31.5) | 32<br>(6.0) | 68 | 468<br>(81.8) | 82<br>(14.3) | 22<br>(3.9) | 9 |
| Washed your hands with soap and water more often than usual | 8,176<br>(95.0) | 371<br>(4.3) | 56<br>(0.7) | 8 | 576<br>(95.7) | 23<br>(3.8) | 3<br>(0.5) | 2 | 561<br>(96.6) | 17<br>(2.9) | 3<br>(0.5) | 0 |
| Kept child(ren) at home before any mandates were put in place | 1,151<br>(53.6) | 957<br>(44.6) | 38<br>(1.8) | 6,465 | 111<br>(58.1) | 75<br>(39.3) | 5<br>(2.6) | 413 | 208<br>(60.6) | 97<br>(28.3) | 38<br>(11.1) | 238 |
| Limiting interactions with people (%) | Y | N | NS | NA | Y | N | NS | NA | Y | N | NS | NA |
| Cancelled or postponed a social event | 8,042<br>(94.8) | 268<br>(3.2) | 178<br>(2.1) | 123 | 580<br>(97.0) | 11<br>(1.8) | 7<br>(1.2) | 6 | 572<br>(98.8) | 6<br>(1.0) | 1<br>(0.2) | 2 |
| Kept away from crowded places | 7,834<br>(92.4) | 367<br>(4.3) | 277<br>(3.3) | 133 | 555<br>(93.8) | 22<br>(3.7) | 15<br>(2.5) | 12 | 575<br>(99.3) | 3<br>(0.5) | 1<br>(0.2) | 2 |
| Tried to avoid people who have the cold or flu-like symptoms | 7,523<br>(89.0) | 514<br>(6.1) | 414<br>(4.9) | 160 | 469<br>(81.1) | 63<br>(10.9) | 46<br>(8.0) | 26 | 515<br>(90.2) | 31<br>(5.4) | 25<br>(4.4) | 10 |
| Avoiding travel (%) | Y | N | NS | NA | Y | N | NS | NA | Y | N | NS | NA |
| Reduced the amount of going to school/university/work | 6,446<br>(88.0) | 839<br>(11.5) | 43<br>(0.6) | 1,283 | 478<br>(84.9) | 81<br>(14.4) | 4<br>(0.7) | 41 | 535<br>(94.4) | 31<br>(5.5) | 1<br>(0.2) | 14 |
| Reduced use of or change the way you use public transport | 5,521<br>(89.6) | 511<br>(8.3) | 127<br>(2.1) | 2,452 | 493<br>(91.3) | 41<br>(7.6) | 6<br>(1.1) | 64 | 562<br>(98.6) | 5<br>(0.9) | 3<br>(0.5) | 11 |
| Reduced the amount you go to shops | 6,888<br>(81.4) | 1,154<br>(13.6) | 421<br>(5.0) | 148 | 401<br>(67.5) | 134<br>(22.6) | 59<br>(9.9) | 10 | 563<br>(97.7) | 8<br>(1.4) | 5<br>(0.9) | 5 |
| Other behaviors not mentioned | 2,094<br>(25.6) | 5,335<br>(65.2) | 760<br>(9.3) | 422 | 166<br>(28.5) | 316<br>(54.2) | 101<br>(17.3) | 21 | 254<br>(44.0) | 274<br>(47.5) | 49<br>(8.5) | 4 |

Y = Yes; N = No; NS = Not sure; NA = Not applicable. For full questionnaire and wording, see Supplement I.
Response percentages may not add up to 100% due to rounding.

**Figure 3.**
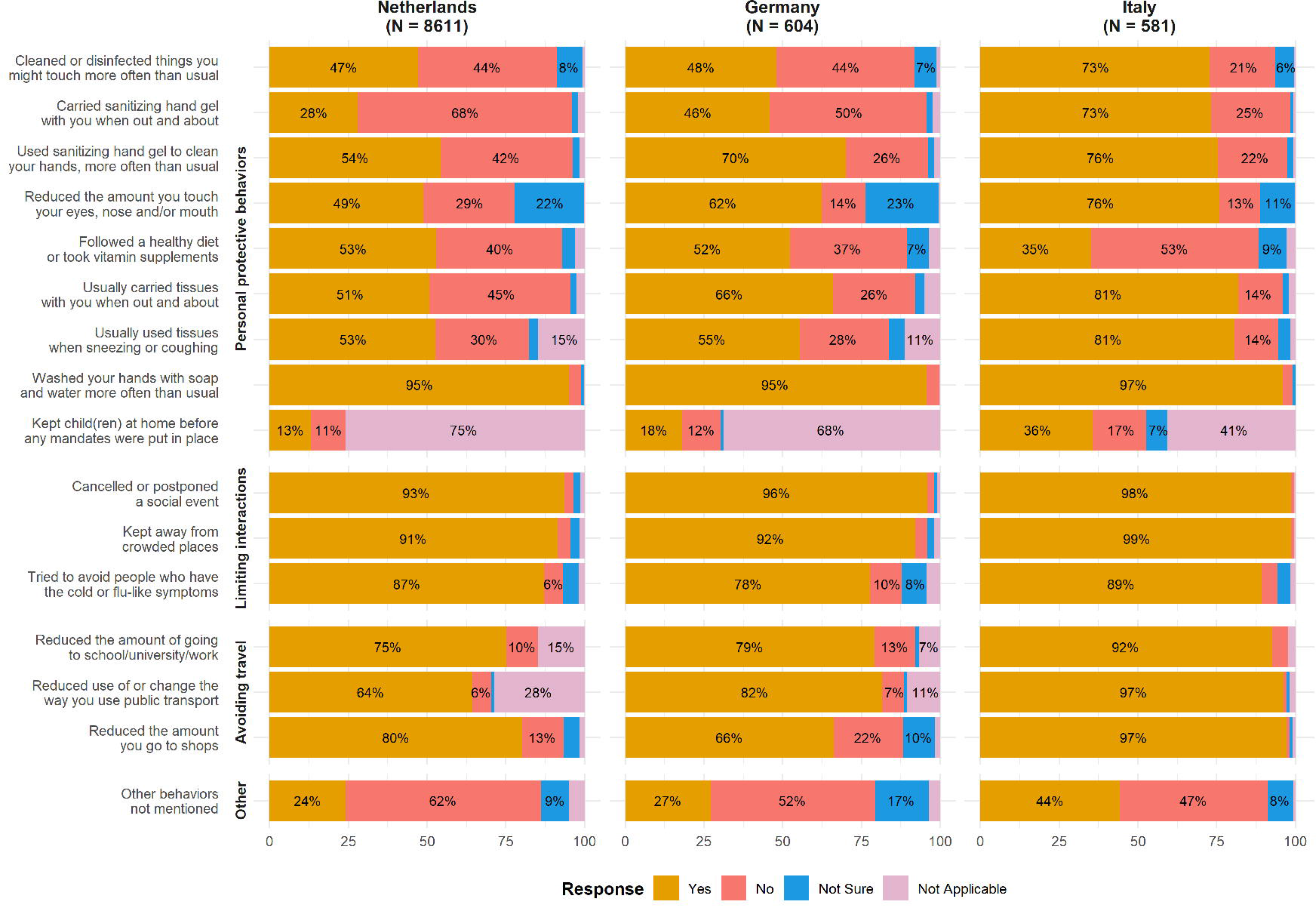
Individual implementation of protective measures in response to COVID-19 pandemic on March 23rd, 2020, by country. Note: Response percentages are rounded and may not add up to 100%. Percentages below 5% omitted.

Behavior related to limiting interactions with people was fairly similar between countries, although respondents from Italy reported more frequently cancelling or postponing social events (98.8%, compared with 94.8% in the Netherlands and 97.0% in Germany) and avoiding crowded places more frequently (99.3%, compared with 92.4% in the Netherlands and 93.8% in Germany). Respondents living in Germany reported avoiding people with cold or flu-like symptoms (81.1%) less frequently than respondents living in Italy (90.2%) or in the Netherlands (89.0%). Regarding behaviors related to avoiding travel, respondents from Italy more often reported to have reduced the amount they went to school or work (94.4%, compared to 88.0% in the Netherlands and 84.9% in Germany, of public transport use (98.6% compared to 89.6% in the Netherlands and 91.3% in Germany), and of going to shops (97.7%, compared to 81.4% in the Netherlands and 67.5% in Germany) (Figure 3 and Table 3). Responses to questions concerning limiting interactions and avoiding traveling may reflect both imposed restrictions and respondents’ awareness and willingness to follow protective measures. Therefore, we additionally asked respondents whether they kept children at home prior to formal mandates to assess the percentage of respondents that applied measures on their own accord. Of those indicating the question was applicable to their situation, in Italy, 60.6% kept their children at home before protective measures compared to 53.6% in the Netherlands and 58.1% in Germany.

### Subgroup analyses among respondents living in the Netherlands

Although we conducted no formal comparisons between the sociodemographic subgroups of participants, some patterns were evident (Supplemental Tables 2a-2r). In general, while there were no meaningful differences in the degree of belief in the effectiveness of protective measures among gender groups, females applied these measures most frequently. Among the different age groups, the belief in the effectiveness of a complete social lockdown differed (e.g. ≤ 20 years: 47.5%, 21-40 years: 62.7%), as well as among subgroups with different daily activities (e.g. retired: 55.4%, homemaker/unemployed: 64.7%) and different education levels (primary/secondary: 54.6%, tertiary academic: 61.1%). Chronically ill patients more frequently reported exhibiting protective measures than respondents without any chronic diseases. Different sociodemographic subgroups used different sources of information to obtain information related to the COVID-19 pandemic. With higher age, the percentage of respondents who agreed they felt sufficiently informed was higher (for example: ≤ 20 years: 87.0%, versus > 60 years: 94.9%)

### Change in responses over time

Immediately following the primary data collection period, we continued collecting data over the next seven days (March 30th, 2020, 11.40 AM, UTC+0) and received responses from 1,588 additional participants, of whom 1,365 reported living in the Netherlands (n=858), in Germany (n=413) and in Italy (n=94). In general, we observed no substantial changes over time (Supplemental Figures 1a-1b), except for a decrease in the belief of effectiveness of a complete social lockdown in Germany (Supplemental Figure 1a). Furthermore, among respondents from the Netherlands, we observed a small increase in the percentages of respondents indicating they believe in the effectiveness of preventive measures (range: 0%- 5%) and those indicating they implemented these measures (range: 0%-10%) across both data collection periods (Supplemental Figures 2a-2b).

## DISCUSSION

Our findings indicate that in three European countries, the Netherlands, Germany, and Italy, the public belief in the effectiveness and the actual implementation of certain protective measures during the ongoing COVID-19 pandemic in March of 2020 was high. Furthermore, residents reported to be sufficiently informed about the ongoing pandemic using various communication channels.

The public belief in the effectiveness of protective measures was highest among respondents residing in Italy, which had the most extensive measures of social lockdown as well as the highest numbers of COVID-19 cases and deaths in Europe in March 2020. Compared to the Netherlands and Germany, respondents living in Italy most often reported not only exhibiting behaviors related to government imposed restrictions but also voluntary hygienic and social measures. Although in general, more than 90% of respondents indicated belief in the effectiveness of imposed measures of social distancing, a complete social lockdown was deemed effective by only 59% of respondents residing in the Netherlands (compared to 77% in Germany and 87% in Italy), where at the time of survey completion, only lighter social distancing measures were enforced. The results of our study suggest that the level of community containment measures implemented by national governments may be rapidly visible in the public beliefs about protective measures, the extent to which people actually exhibit these relevant behaviors, and reflect the severity of the outbreak situation in a given country.

To the best of our knowledge, to date, few data on this topic are available. Results from two recently published survey studies conducted in the USA, the UK, and China primarily focus on the respondents’ knowledge about COVID-19 and assess understanding pertaining to the disease course[13,14]. Furthermore, another study conducted between January 24th and February 13th, 2020 among 1715 Hong Kong residents showed that most respondents obtained information on the COVID-19 pandemic from social media and websites[15]. We found that traditional information sources (e.g. television and news) were used most frequently among our respondents. Our study further corroborates and adds to these first findings with similar results regarding beliefs in the effectiveness of hygienic and social distancing measures and the extent to which these measures were exhibited in a European study population.

## Social distancing and other behavioral measures

Individuals’ adherence to country-specific mitigation measures has the potential to influence the course of the COVID-19 pandemic. Social (physical) distancing has been proposed as one of the most effective measures for mitigating pandemics caused by viruses, including COVID-19[4,7,16]. Large-scale simulation studies have found that closure of borders is only effective to prevent further spread of the virus if they are implemented for more than two weeks and prevent international travel [8]. Moreover, the peak attack rate can be decreased by case isolation, household quarantine, and school, university and work closure[8].

As the transmissibility of the SARS-CoV-2 virus is estimated to be similar to or higher than previous coronaviruses such as SARS, social isolation measures are particularly important [17,18]. In the current COVID-19 pandemic, models have shown that the Wuhan quarantine reduced transmission of COVID-19 cases from mainland China to other countries by 77% by early February [9]. Besides social distancing, other behavioral protective measures have also been deemed effective in the mitigation of the current pandemic. For instance, regular hand washing may result in a reduction of peak infection rate up to 65% with a delay of 2.7 months and a 29% decrease in total infection rate[19]. Generally, preventive, precautionary behavior is more commonly observed, in females, and in older persons [20–22], which was also reflected in our findings from Europe during the ongoing COVID-19 pandemic.

## Provision and acquirement of information during pandemics

Transparent, timely, and easy-to-understand information is essential to increase trust in national governments during pandemics[23]. The increasing use of portable devices and social media is evident in our findings, which indicate frequent use of social media to acquire pandemic-related information (range across countries: 40.0%-55.8%). However, in recent epidemics and pandemics, a substantial amount of online information, especially distributed via social media, was found to be incorrect and misleading[24,25]. Environmental cues to follow behavioral recommendations, favorable attitudes towards prevention measures, and knowledge about the virus were associated with exhibiting protective behavior[5]. Therefore accurate information provision via social media channels is crucial, besides information via traditional information sources.

## Study strengths and limitations

Given the evolving pandemic situation, we felt it was important to develop, translate, and disseminate our questionnaire rapidly to capture a snapshot of public perceptions and behaviors in ‘real time’ as the COVID-19 crisis unfolded in Europe and as policy makers enacted formal containment measures in several European countries. Many items in our survey were adapted from an existing validated questionnaire created to assess perceptions and behaviors in response to influenza[10]. We attempted to make our survey accessible to participants of diverse backgrounds by providing the survey in eight languages. These translations could be readily adapted for use in future studies on other viral pandemics.

Readers should consider some important limitations when interpreting our findings. First, since the survey was web-based and recruitment was largely through digital channels including social media, we acknowledge the potential for selection bias. We cannot assume that our study population is representative for the individual countries and acknowledge possible over-representation of health-conscious individuals and those more informed or concerned about the outbreak. However, under the exceptional current circumstances, members of the general public who normally would not participate in health-related surveys may be more likely to participate given the media attention, severity, and the outbreak’s large impact on many aspects of daily life. Furthermore, with the social (physical) distancing recommendations and enforced measures in place, many confined to their homes have turned to social media and other web-based platforms for social communication, including those belonging to the older age groups.

Second, the number of completed survey responses was much higher among residents of the Netherlands compared to Italy or Germany. Since the majority of our research team members are based in the Netherlands and the largest dissemination efforts occurred there, this is not surprising. Third, while the governments of Germany, Italy, and the Netherlands enacted different mitigation measures in each country, our survey was not adapted to country-specific nuances. Hence, we acknowledge that our results might not fully depict whether residents of these countries actually exhibit their country-specific measures. Fourth, we cannot rule out that an individual completed the survey more than once on multiple devices, in another browser, or by clearing cookies; however, repeat submissions from the same device were not accepted.

Fifth, with regard to the secondary analysis over the extended data collection period, we observed no major changes in the aggregated answers over time; however, we acknowledge a possible delay between the implementation of formal community isolation measures and the subsequent information uptake and application of these measures by residents. During the extended data collection period, response rates were lower than in the primary collection period, especially from respondents living in Italy.

Finally, we emphasize the aim of our study was descriptive, and we caution readers against drawing causal conclusions regarding the observed differences. A formal comparison between countries would require appropriate analytical consideration of variables taking into account sociocultural (including educational systems), political, and structural contexts in each country. In an effort to help the reader better understand each country-specific setting we included information about the stage of containment measures enacted within the included countries in a timeline encapsulating the data collection period.

## CONCLUSION

The extent to which individuals internalize and respond to (government-mandated) mitigation measures and recommendations is critical to the control of the spread of the SARS-CoV-2 virus and to optimize outcomes during the current COVID-19 pandemic. In our survey study of the general public living in the Netherlands, Germany, and Italy, we found that approval and application of publicly enforced and self-initiated protective measures were highest in Italy, the region with the most extensive measures of social lockdown and highest burden (number of cases and deaths) in Europe, during the study time period in mid-March, 2020. Media channels used to acquire information and the extent to which respondents felt sufficiently informed about the COVID-19 pandemic differed per country and among sociodemographic subgroups in the Netherlands. No substantial changes in the perceived effectiveness of behavioral protective measures and the implementation of these measures in these countries were observed between March 19th and March 30th, 2020, as the COVID-19 pandemic continued to evolve in Europe and formal community isolation measures became stricter. We believe these insights are valuable to inform the information dissemination and infection control strategies of governments and public health organizations during the current crisis and also for future pandemics.

## Summary boxes

### What is already known on this topic

- Social distancing and hygienic measures are effective in mitigating pandemics such as the COVID-19 pandemic.
- Public beliefs regarding the effectiveness of protective measures, and the extent to which these measures have been applied by European residents during the COVID-19 pandemic remain unknown.

### What this study adds

- In three European countries, the degree of public belief in the effectiveness of protective measures was high and respondents living in the Netherlands, Germany and Italy reported feeling sufficiently informed by various communication channels.
- Implementation of enacted and self-initiated measures differed between countries and were highest among respondents living in Italy, who were subjected to the most elaborate measures of social lockdown and experienced the greatest outbreak burden.

## Data Availability

Requests for data sharing can be directed to AHZN. All survey items are included as supplemental material.
Upon request, the data will be available for policy makers and government bodies.

## Contributors

AHZN conceived this study and AHZN, KM, and JLR developed the study design. All authors contributed to design the survey, translation and verification of the survey items in German, Dutch or Italian, and distribution of the survey insocial and professional networks. KM, TG, MP, and RM conducted the statistical analysis and data visualization. MCG and ATCJ classified countries on containment stage. KM, JLR and AHZN drafted the manuscript with support from MM, KA, ATCJ, TG and MP. The COVID-19 study group contributed to translation, pilot testing of the survey, dissemination, and country community isolation measure classification. All authors and study group members helped interpret the results and critically revised the final manuscript. AHZN supervised the project. The corresponding author attests that all listed authors meet authorship criteria and that no others meeting the criteria have been omitted.

## Ethical approval

The study was reviewed and granted exempted status from medical ethical approval by the Institutional Review Board of Leiden University Medical Center in The Netherlands (protocol number: N20-037).

## Data sharing

Requests for data sharing can be directed to AHZN. All survey items are included as supplemental material.

## Transparency

The corresponding author (AHZN) affirms that this manuscript is an honest, accurate, and transparent account of the study being reported; that no important aspects of the study have been omitted; and that any discrepancies from the study as planned have been explained.

## Patient and Public Involvement

The target population, the general public, was actively involved in the dissemination of this survey. No patients were involved in this study, as the target group was the general public.

## Dissemination declaration

Upon request, the data will be available for policy makers and government bodies.

## Acknowledgements

*Ilaria Vitulano*

*Büşra Nur Bozok*

*María Alcántara Laguna*

*Ariane Scholten*

*Renate Meier*

*Shabbir R Shahnahpur*

*Marcin M Fiszer*

